# TruBlk™ Shilajit Resin Improves Sexual Function in Men with Erectile Dysfunction: A Randomised, Double-Blind, Placebo-Controlled, Multicenter Trial

**DOI:** 10.64898/2026.10.23.26366294

**Authors:** Divya Yadav, Utsa Basu, Kurmi Prakash Harishchandra, Manoj Karwa, Surya Teja Reddy, Amandeep Singh, Karan Shah

## Abstract

Erectile dysfunction and age-related testosterone decline are common and growing concerns among ageing men, yet current pharmacological options are constrained by tolerability, cost, and reluctance to start hormone-based therapy, leaving a real gap for safe, long-term alternatives. Shilajit, a resinous plant exudate long used in Ayurvedic medicine, is increasingly proposed to fill this gap with its bioactive constituents thought to confer adaptogenic, antioxidant, and androgen-modulating effects. However, clinical support for this use remains thin, with most existing trials small and conducted in healthy men rather than those with diagnosed erectile dysfunction.

In this randomised, double-blind, placebo-controlled, multicenter trial (CTRI/2025/07/090919), 100 men aged 35–55 years with mild-to-moderate erectile dysfunction (IIEF erectile function domain score 11– 25) and symptoms suggestive of androgen deficiency were enrolled across two sites. Participants received 250 mg twice daily of standardised TruBlk™ Shilajit resin or matched placebo for 90 days. Primary endpoints, as registered, were change from baseline in IIEF total and domain scores, total and free testosterone and cortisol, and Hypogonadism Impact of Symptoms Questionnaire (HIS-Q) domain scores. Secondary endpoints included nitric oxide, Fatigue Severity Scale (FSS), and Participant and Investigator Global Assessments. Semen parameters were assessed in a subset of participants.

IIEF total score improved significantly more in the TruBlk™ group (+32.1% vs. +13.6%; d=0.67, p=0.005), with significant gains in four of the five domains, namely erectile function, orgasmic function, overall satisfaction, and intercourse satisfaction (d=0.52–0.61, p=0.011–0.044), while sexual desire showed a smaller, non-significant improvement (d=0.44, p=0.109). This was accompanied by significant increases in total testosterone (+46.1% vs. +10.0%; d=0.44, p=0.030) and free testosterone (+37.4% vs. +6.8%; d=0.41, p=0.048). HIS-Q symptom domains showed a similar pattern to IIEF, with Day 90 differences favouring TruBlk™ in three of five domains (sexual, energy, and sleep; d=−0.59 to −0.82, p<0.02), while cognition and mood showed smaller, non-significant differences (d=−0.46 to −0.49, p=0.06–0.10). Among secondary endpoints, fatigue fell substantially more with TruBlk™ than with placebo, with FSS scores decreasing by 51.8% vs. 27.4% from baseline, a large between-group effect at Day 90 (d=−1.13, p<0.0001). At Day 90, 56–58% of TruBlk™ participants were rated “Much” or “Very Much Improved” on global assessment, compared with 12–14% of placebo. Nitric oxide showed no significant between-group difference. In an exploratory semen-analysis subset (n=57), sperm concentration increased with TruBlk™ relative to placebo (δ=0.53, p<0.001), and in a subset of 40 participants assessed for motility, total motility (δ=0.63, p<0.001) and progressive motility (d=0.82, p=0.014) also improved.

Ninety days of standardised TruBlk™ Shilajit resin improved erectile function and increased total and free testosterone in men with mild-to-moderate erectile dysfunction, with improvement in several hypogonadism-related symptom domains, reduced fatigue and, in an exploratory subset, improved sperm concentration and motility. These results support standardised Shilajit resin as a promising intervention for sexual dysfunction in a clinically diagnosed population, meriting further clinical evaluation.

## 1. Introduction

Male sexual dysfunction and age-related testosterone decline are increasingly recognised as significant public health concerns, with substantial implications for quality of life, psychological wellbeing, and relationship satisfaction in ageing men. Erectile dysfunction alone is projected to affect over 300 million men worldwide, with prevalence rising sharply from under 10% in men below 40 to more than half of men over 70 [1, 2]. Testosterone follows a comparable age-related trajectory, with a growing proportion of ageing men affected by clinically low levels [3]. Despite the scale of this burden, pharmacological options remain limited by tolerability concerns, cost, and patient reluctance to initiate hormone-based therapies, creating sustained interest in safe, natural alternatives that can be used long-term.

Shilajit is a naturally occurring resinous exudate, formed through the slow humification of plant matter compressed within high-altitude rock strata, with centuries of documented use in Ayurvedic medicine as a *rasayana* (rejuvenative) agent. Its principal bioactive constituents include fulvic acid, which functions as a carrier and chaperone molecule facilitating cellular uptake of trace minerals and other bioactives; and shilarathenes™, a broader spectrum of dibenzo-α-pyrones, urolithin-associated metabolites, phenolic acids, and flavonoids proposed to underlie its adaptogenic, antioxidant, and androgen-modulating properties [4-7].

Shilajit has attracted growing attention in the sexual wellness and men’s health space in recent years, driven in part by controlled trials reporting favourable shifts in total and free testosterone in healthy middle-aged men, alongside preliminary evidence suggesting benefits for broader measures of sexual health [8]. However, this attention has outpaced the clinical evidence base. An added confounder across much of this literature is the lack of standardised, quality-controlled shilajit ingredients in the market; while a small number of established suppliers have demonstrated that consistent, verified bioactive content is achievable, the majority of commercially available shilajit remains unstandardised, and much of the existing clinical evidence is concentrated on a single such extract. Furthermore, existing controlled trials remain small, single-site, and conducted predominantly in healthy volunteers rather than in men with diagnosed sexual dysfunction.

This trial uses TruBlk™, a shilajit extract standardised through HPLC-validated quantification to guarantee a minimum of 60% fulvic acid and 10% shilarathenes™ by weight, providing a reproducible, authenticated bioactive profile that unstandardised commercial resins cannot guarantee [9]. The present study addresses this gap through a randomised, double-blind, placebo-controlled, multicenter trial designed to evaluate standardised TruBlk™ Shilajit resin in men aged 35 to 55 years with mild-to-moderate erectile dysfunction (IIEF erectile function domain score 11–25) and symptoms suggestive of androgen deficiency (positive ADAM questionnaire), with the aim of establishing whether shilajit’s effects extend beyond the healthy-volunteer populations studied to date into a diagnosed, clinically relevant target group.

## 2. Methods

### 2.1 Study Design

This was a randomised, double-blind, placebo-controlled, parallel-group, multicenter trial conducted at two sites in India between September 2025 and January 2026. The trial was conducted in accordance with ICH-GCP guidelines and the Declaration of Helsinki, and approval was obtained from the Institutional Ethics Committees of both sites prior to commencement. All participants provided written informed consent before enrolment. The trial was prospectively registered at Clinical Trials Registry India (CTRI/2025/07/090919) prior to enrolment of the first participant. This manuscript was prepared in accordance with the CONSORT 2010 guidelines for reporting randomised trials [10].

### 2.2 Participants

Males aged 35 to 55 years with mild-to-moderate erectile dysfunction, defined as an IIEF erectile function domain score of 11–25 [11], and a positive Androgen Deficiency in Aging Males (ADAM) questionnaire (≥3 positive responses), were enrolled [12]. Eligible participants had a BMI of 30 kg/m² or below, were willing to abstain from other sexual health supplements for the duration of the study, and were capable of providing written informed consent.

Individuals were excluded if they had severe erectile dysfunction (IIEF-EF score below 11) or a known organic cause of erectile dysfunction [13], such as Peyronie’s disease or penile trauma. Further exclusion criteria included current use of PDE5 inhibitors or testosterone replacement therapy; a history of prostate cancer or PSA above 4 ng/mL; uncontrolled diabetes (HbA1c above 8%), hypertension, or cardiovascular disease; hepatic or renal impairment (ALT/AST greater than twice the upper limit of normal, or serum creatinine above 1.5 mg/dL); a history of psychiatric disorders requiring medication; known allergy or hypersensitivity to Shilajit or its components; and participation in another clinical trial within the preceding 30 days.

### 2.3 Randomisation and Blinding

Eligible participants were randomised 1:1 to receive either TruBlk™ Shilajit Resin or matching placebo, using a computer-generated randomisation sequence with block sizes of four, stratified by study site. Allocation concealment was maintained using sequentially numbered, sealed, opaque envelopes. Participants, investigators, outcome assessors, and data analysts remained blinded to treatment allocation throughout the study, with investigational product and placebo identical in appearance, taste, and packaging.

### 2.4 Intervention

Participants in the active group received TruBlk™ Shilajit Resin, a purified and standardised extract manufactured by Amaara Botanicals Pvt Ltd. and standardised to ≥60% fulvic acid and ≥10% shilarathenes™, administered orally at 250 mg twice daily (morning and evening, with meals) for 90 days. Placebo sachets containing inert excipients and identical in appearance, taste, and packaging, were administered on the same schedule. Participants were instructed to maintain their usual diet and lifestyle throughout the study and to record daily compliance in a diary; unused investigational product was returned at each visit for compliance assessment.

### 2.5 Outcome Measures

Participants were assessed at screening (Day −14 to Day 0), baseline (Day 0), and follow-up visits on Day 30, Day 60, and Day 90 (±3 days), with the Day 90 visit comprising the full outcome and safety battery alongside return of unused medication.

The primary endpoints, as registered (CTRI/2025/07/090919), were change from baseline in IIEF total score and the five IIEF domain scores (erectile function, orgasmic function, sexual desire, intercourse satisfaction, overall satisfaction; in serum total testosterone, free testosterone (Day 90); and in Hypogonadism Impact of Symptoms Questionnaire (HIS-Q) domain scores [14]. Secondary endpoints comprised change from baseline in serum nitric oxide, Fatigue Severity Scale (FSS) [15], and Participant and Investigator Global Assessments (PGA, IGA). Total testosterone and free testosterone were measured by chemiluminescence immunoassay (CLIA) from morning fasting blood samples; nitric oxide was measured using the Griess reaction method.

Semen analysis was performed at baseline and Day 90 in the subset of participants who consented (n = 57 across both sites), within one hour of collection and according to WHO 2010 guidelines. Sperm concentration and motility, graded as rapid progressive, slow progressive, non-progressive, or immotile, are reported here. Morphology (Kruger strict criteria) and vitality (eosin-nigrosin staining) were recorded at Site 2 only and are not reported [16].

Safety endpoints included adverse events and serious adverse events, vital signs (blood pressure, heart rate, respiratory rate), anthropometric measurements (weight, BMI), and clinical laboratory parameters (haematology, biochemistry, liver and kidney function), assessed at each visit.

### 2.6 Statistical Analyses

The outcomes assessed at four timepoints (IIEF total score, IIEF domain scores, FSS, and HIS-Q domain scores) were analysed using a mixed-effects model (REML) with treatment as a between-subjects factor and time as a within-subjects factor, without assuming sphericity; the treatment × time interaction was the primary test of differential response between groups, and between-group differences at Day 90 were assessed by pairwise comparison, Sidak-corrected for the four study visits.

The outcomes assessed at two timepoints (total and free testosterone, nitric oxide, cortisol, and semen parameters) were compared by between-group analysis for change from baseline. Normality of baseline values and of change scores was each assessed independently by Shapiro–Wilk test: normally distributed variables were compared by unpaired t-test and are presented as mean ± SD, with Cohen’s d as the effect size; non-normally distributed variables were compared by Mann-Whitney U test and are presented as medians with interquartile ranges, with Cliff’s delta as the effect size and the Hodges– Lehmann estimate (95% CI) as the between-group difference.

Sperm concentration was analysed in participants with paired semen samples from both sites (TruBlk™ n = 29, placebo n = 28), and motility in participants with paired samples from Site 2 (TruBlk™ n = 20, placebo n = 20); semen analyses were exploratory. Within-group changes were assessed by paired t-test or Wilcoxon signed-rank test as appropriate, and values below the assay detection limit were imputed at the detection limit. Participant and Investigator Global Assessments were assessed as a single post-treatment rating at Day 90 and compared between groups by Mann-Whitney U test on the seven-point ordinal grade.

Analyses were performed using GraphPad Prism version 10, except for Cliff’s delta, which was computed externally. A two-sided p-value below 0.05 was considered statistically significant, except where otherwise noted for Sidak-corrected comparisons. The sample size of 100 participants (50 per group) was chosen to match a concurrently conducted trial of identical design and dosing [9]. With 50 participants per group and a two-sided α of 0.05, the study had 80% power to detect a standardised between-group difference of d = 0.57, and 90% power to detect d = 0.66.

## 3. Results

### 3.1 Participant Flow and Baseline Characteristics

One hundred and two participants were screened for eligibility, of whom 2 did not meet inclusion criteria and were excluded prior to randomisation. The remaining 100 were randomised equally to TruBlk™ (n=50) or placebo (n=50), stratified by site (Site 1: n=45; Site 2: n=55). All 100 participants received their allocated treatment, and none were lost to follow-up or discontinued the intervention, giving a trial completion rate of 100%. All randomised participants were included in the full analysis set (TruBlk™ n=50, placebo n=50). Full compliance with the supplementation schedule was recorded in both groups. Semen analysis was performed in a consenting subset (TruBlk™ n=29, placebo n=28), with a subset of patients who hadmotility assessed (n=20 per group). Participant flow is summarised in Figure 1.

**Figure 1.**
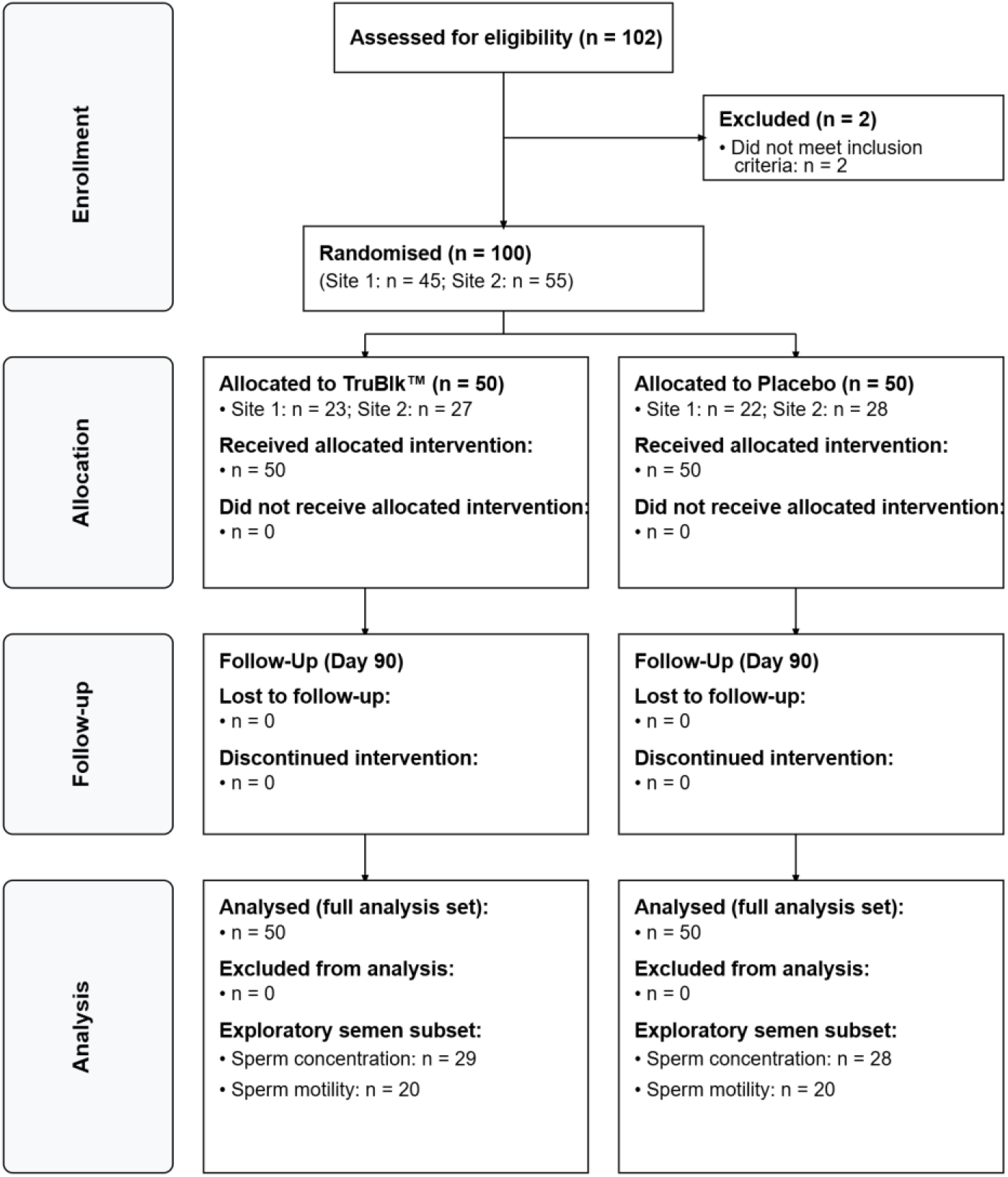
Consort Diagram. Flow of participants through each stage of the randomised, double-blind, placebo-controlled trial of TruBlk™ (CTRI/2025/07/090919). Of 102 individuals assessed for eligibility, 2 were excluded for not meeting inclusion criteria, yielding 100 participants who were randomised 1:1, stratified by site, to receive TruBlk™ 250 mg twice daily (n = 50) or matching placebo (n = 50) for 90 days. No participants were lost to follow-up or discontinued the intervention in either group, and all 100 were included in the full analysis set.

Groups were well matched for baseline clinical and biochemical characteristics, with one exception: participants in the Active group were significantly older than those in the Placebo group (median 47.0 years, IQR 42.0–51.0, vs. 43.50 years, IQR 40.0–47.75; Mann-Whitney U, p=0.31). BMI was comparable between groups (median 25.62 kg/m², IQR 23.11–27.11, vs. 26.12 kg/m², IQR 24.00– 27.53; p=0.430), as was the distribution of participants across the two study sites (Fisher’s exact, p=1.00). Baseline values for the primary outcomes were comparable between groups as were IIEF erectile function domain score, ADAM total score, total and free testosterone, nitric oxide, HIS-Q domain scores, and FSS score (all p>0.05). Baseline characteristics are presented in Table 1.

**Table 1.** Baseline Characteristics of Participants.

| **Variable** | **Active (n=50)** | **Placebo (n=50)** | **Test** | **p-value** |
|---|---|---|---|---|
| Age (years) | 47.00 [42.00–51.00] | 44.50 [40.00–47.75] | Mann-Whitney U | 0.31 |
| BMI (kg/m²) | 25.62 [23.11–27.11] | 26.12 [24.00–27.53] | Mann-Whitney U | 0.430 |
| Site distribution, n (%) | Site 1: 23, Site 2: 27 | Site 1: 22, Site 2: 28 | Fisher's exact | 1.000 |
| ADAM Total Score | 6.00 [6.00–7.00] | 6.00 [6.00–7.00] | Mann-Whitney U | 0.861 |
| IIEF Total Score | 24.00 [21.00–39.00] | 23.00 [22.00–40.75] | Mann-Whitney U | 0.582 |
| IIEF-EF Domain Score | 12.00 [11.00–16.75] | 13.00 [11.00–16.75] | Mann-Whitney U | 0.669 |
| Total Testosterone (ng/dL) | 385.74 [14.81–538.11] | 321.95 [16.30–485.94] | Mann-Whitney U | 0.758 |
| Free Testosterone (pg/mL) | 15.50 [1.19–21.40] | 13.30 [1.88–19.90] | Mann-Whitney U | 0.603 |
| FSS | 50.00 [44.00–53.00] | 51.00 [43.25–52.75] | Mann-Whitney U | 0.58 |
| Nitric Oxide (µmol/L) | 24.55 [18.36–37.47] | 23.48 [13.23–32.35] | Mann-Whitney U | 0.157 |

### 3.2 Erectile Function (IIEF)

At baseline, groups were well matched for IIEF total score, with no significant difference between those receiving TruBlk™ (30.00 ± 9.53) and placebo (30.28 ± 9.37; unpaired t-test, p = 0.883). Over the 90-day study period, IIEF total score increased significantly in both groups (main effect of time: F(1.48, 145.51) = 193.53, p < 0.0001), consistent with a general improvement in erectile function across the cohort. The rate of improvement differed between groups, with a steeper trajectory in the TruBlk™ group producing a significant treatment × time interaction (F(1.48, 145.51) = 35.92, p < 0.0001), indicating that the two groups diverged rather than improving in parallel. By day 90, participants receiving TruBlk™ had gained 9.64 points (+32.1%) compared with 4.12 points (+13.6%) in the placebo group (difference in mean change 5.52 points). At Day 90, IIEF total score was 5.24 points higher with TruBlk™ than placebo (39.64 vs. 34.40; 95% CI 2.12 to 8.36), a moderate effect (Cohen’s d = 0.67, t(98) = 3.33, Sidak-corrected p = 0.005) (Figure 2A).

**Figure 2.**
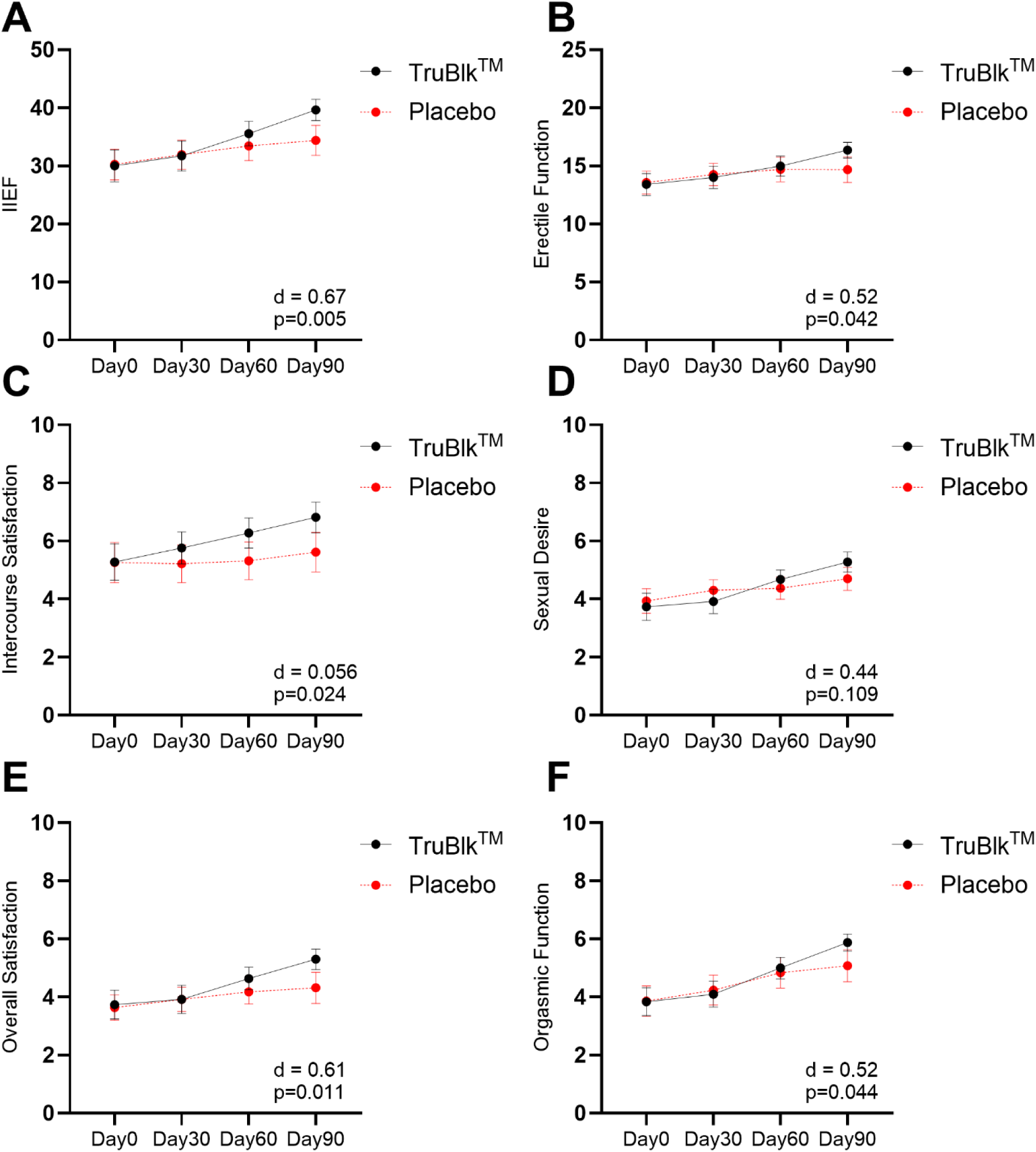
IIEF total score and domain scores over 90 days. (A) IIEF total score, (B) erectile function, (C) intercourse satisfaction, (D) sexual desire, (E) overall satisfaction, and (F) orgasmic function domain scores, assessed at baseline (Day 0) and at days 30, 60, and 90 in participants receiving TruBlk™ (black, n = 50) or placebo (red, n = 50). Data are mean ± SEM.

Individual IIEF domain scores showed a treatment × time interaction favouring the TruBlk™ group in four of the five domains: erectile function (F(1.84, 180.49) = 19.73, p < 0.0001; +2.96 vs. +1.10 points, Day 90; Cohen’s d = 0.52, Sidak p = 0.042), intercourse satisfaction (F(2.08, 203.37) = 17.75, p < 0.0001; +1.54 vs. +0.36; d = 0.56, Sidak p = 0.024), overall satisfaction (F(1.64, 160.51) = 8.36, p < 0.001; +1.56 vs. +0.68; d = 0.61, Sidak p = 0.011), and orgasmic function (F(1.67, 163.60) = 6.22, p = 0.004; +2.04 vs. +1.22; d = 0.52, Sidak p = 0.044). Sexual desire showed a significant omnibus interaction across the full 90-day trajectory (F(2.45, 239.86) = 11.24, p < 0.0001), but the between-group difference at Day 90 specifically did not reach significance after correction for multiple comparisons (+1.54 vs. +0.76 points; d = 0.44, Sidak p = 0.109). All five domains were well matched at baseline (all p > 0.5) (Table 2, Figure 2B-F).

**Table 2.** IIEF Total Score and Domain Scores at Baseline and Day 90.

| Endpoint | Group | Baseline | Day 90 | Change | Effect size | p |
|---|---|---|---|---|---|---|
| IIEF Total Score | TruBlk™ | 30.00 ± 9.53 | 39.64 ± 6.47 | +9.64 | d = 0.67 | 0.005 |
|  | Placebo | 30.28 ± 9.37 | 34.40 ± 9.04 | +4.12 |  |  |
| Erectile Function | TruBlk™ | 13.40 ± 3.33 | 16.36 ± 2.38 | +2.96 | d = 0.52 | 0.042 |
|  | Placebo | 13.58 ± 3.44 | 14.68 ± 3.89 | +1.10 |  |  |
| Orgasmic Function | TruBlk™ | 3.84 ± 1.68 | 5.88 ± 1.02 | +2.04 | d = 0.52 | 0.044 |
|  | Placebo | 3.86 ± 1.83 | 5.08 ± 1.94 | +1.22 |  |  |
| Sexual Desire | TruBlk™ | 3.74 ± 1.64 | 5.28 ± 1.20 | +1.54 | d = 0.44 | 0.109 |
|  | Placebo | 3.94 ± 1.49 | 4.70 ± 1.40 | +0.76 |  |  |
| Overall Satisfaction | TruBlk™ | 3.74 ± 1.75 | 5.30 ± 1.25 | +1.56 | d = 0.61 | 0.011 |
|  | Placebo | 3.64 ± 1.52 | 4.32 ± 1.89 | +0.68 |  |  |
| Intercourse Satisfaction | TruBlk™ | 5.28 ± 2.19 | 6.82 ± 1.86 | +1.54 | d = 0.56 | 0.024 |
|  | Placebo | 5.26 ± 2.44 | 5.62 ± 2.39 | +0.36 |  |  |

### 3.3 Hormonal Parameters

Total testosterone was comparable between groups at baseline (median 385.74 [IQR 14.81–538.11] ng/dL in the TruBlk™ group vs. 321.95 [16.30–485.94] ng/dL in placebo; p = 0.758). Over the 90-day period, total testosterone increased significantly in the TruBlk™ group (+155.27 ± 294.27 ng/dL, +46.1%; within-group p = 0.0006) but remained unchanged in placebo (+32.62 ± 259.70 ng/dL, +10.0%; p = 0.379). This produced a significant between-group difference of 122.65 ng/dL (95% CI 11.86 to 233.44), representing a small-to-moderate effect (Cohen’s d = 0.44, p = 0.030) (Figure 3A).

**Figure 3.**
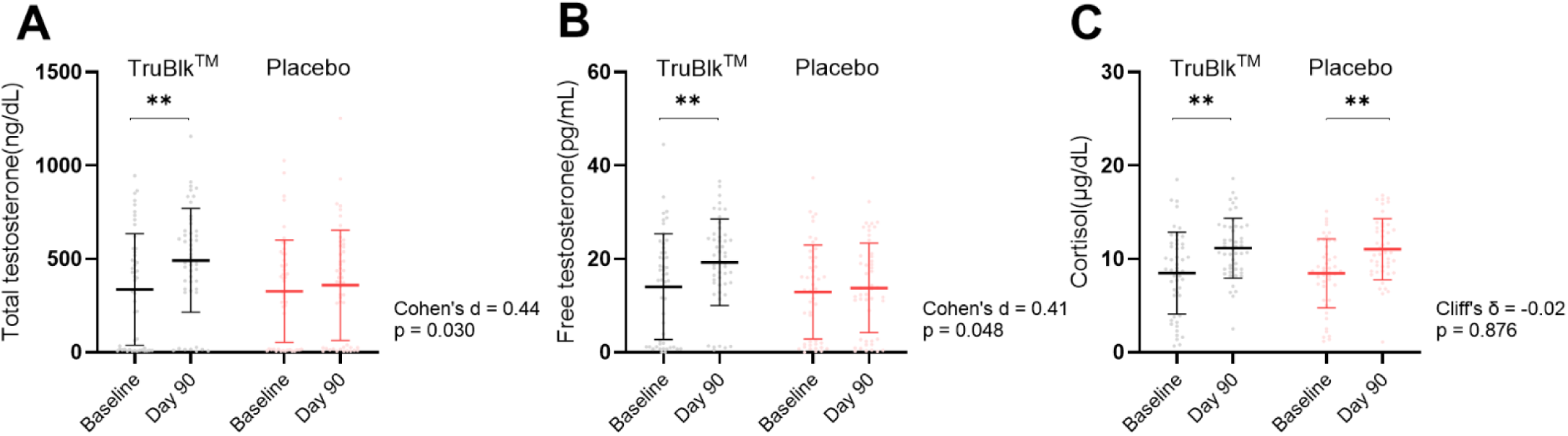
Effect of TruBlk™ Shilajit on hormonal parameters. (A) Total testosterone, (B) free testosterone, and (C) cortisol in participants receiving TruBlk™ (black) or placebo (red) at baseline and day 90. Data are shown as mean ± SD. Asterisks denote within-group change from baseline (**p < 0.01); Cohen’s d (A, B) and Cliff’s δ (C) denote the effect size for the between-group difference in change from baseline, with the corresponding p value.

Free testosterone was likewise comparable between groups at baseline (median 15.50 [IQR 1.19– 21.40] pg/mL in the TruBlk™ group vs. 13.30 [1.88–19.90] pg/mL in placebo; p = 0.603). Over the 90-day period, free testosterone increased significantly in the TruBlk™ group (+5.25 ± 11.29 pg/mL, +37.4%; within-group p = 0.002) but remained unchanged in placebo (+0.88 ± 10.00 pg/mL, +6.8%; p = 0.551). This produced a significant between-group difference of 4.37 pg/mL (95% CI 0.05 to 8.69), representing a similar small-to-moderate effect (Cohen’s d = 0.41, p = 0.048) (Figure 3B).

Cortisol was comparable between groups at baseline (8.48 ± 4.40 µg/dL in the TruBlk™ group vs. 8.45 ± 3.68 µg/dL in placebo; p = 0.970). Over the 90-day period, cortisol increased in both groups to a similar degree (TruBlk™: +2.68 ± 5.80 µg/dL, +31.5%, within-group p = 0.0073; placebo: +2.58 ± 5.36 µg/dL, +30.6%, within-group p = 0.0021), with no between-group difference in change from baseline (mean difference 0.09 µg/dL; Cliff’s δ = −0.02, p = 0.876) (Figure 3C).

### 3.4 Hypogonadism-Related Symptoms (HIS-Q)

Groups were well matched at baseline across all five HIS-Q domains (all p>0.44). Over the 90-day period, symptom scores decreased in both groups, with a treatment × time interaction favouring the TruBlk™ group in four of the five domains: Sexual (F=9.97, p<0.001), Energy (F=6.34, p=0.004), Cognition (F=3.76, p=0.037), and Mood (F=4.37, p=0.026). Sleep showed an interaction that did not reach significance (F=3.35, p=0.051). By Day 90, the TruBlk™ group showed numerically lower (less symptomatic) scores than placebo across all five domains: Sexual (1.31 ± 0.53 vs. 1.75 ± 0.55), Energy (1.15 ± 0.53 vs. 1.51 ± 0.58), Sleep (1.57 ± 0.58 vs. 1.89 ± 0.48), Cognition (1.37 ± 0.65 vs. 1.70 ± 0.67), and Mood (1.44 ± 0.66 vs. 1.73 ± 0.63), with effect sizes ranging from small to large (Cohen’s d = −0.46 to −0.82). Sidak-corrected comparisons at Day 90 were significant for Sexual, Energy, and Sleep (all p<0.02), but not for Cognition (p=0.060) or Mood (p=0.096) (Table 3).

**Table 3.** HIS-Q Domain Scores (0–4 scale; lower = less symptomatic)

| **Domain** | **Baseline (TruBlk™ vs. Placebo)** | **Interaction** | **Day 90 (TruBlk™ vs. Placebo)** | **Cohen's d** | **Sidak p** |
|---|---|---|---|---|---|
| Sexual | 2.37±0.33 vs. 2.32±0.38, p=0.521 | F=9.97, p<0.001 | 1.31±0.53 vs. 1.75±0.55 | −0.82 | <0.001 |
| Energy | 2.40±0.83 vs. 2.29±0.69, p=0.485 | F=6.34, p=0.004 | 1.15±0.53 vs. 1.51±0.58 | −0.64 | 0.008 |
| Sleep | 2.15±0.56 vs. 2.21±0.56, p=0.636 | F=3.35, p=0.051 | 1.57±0.58 vs. 1.89±0.48 | −0.59 | 0.017 |
| Cognition | 2.27±0.40 vs. 2.33±0.38, p=0.444 | F=3.76, p=0.037 | 1.37±0.65 vs. 1.70±0.67 | −0.49 | 0.060 |
| Mood | 2.31±0.36 vs. 2.26±0.35, p=0.447 | F=4.37, p=0.026 | 1.44±0.66 vs. 1.73±0.63 | −0.46 | 0.096 |

### 3.5 Secondary Endpoints

#### 3.5.1 Fatigue Severity Scale

At baseline, groups were well matched for Fatigue Severity Scale, with no significant difference between those receiving TruBlk™ (48.78 ± 5.10) and placebo (48.34 ± 5.17; unpaired t-test, p = 0.669). Over the 90-day study period, FSS decreased significantly in both groups (main effect of time: F(1.25, 122.18) = 251.38, p < 0.0001), reflecting a general reduction in fatigue across the cohort. However, the rate of improvement differed markedly between groups, with a steeper decline in the TruBlk™ group producing a significant treatment × time interaction (F(1.25, 122.18) = 27.82, p < 0.0001), indicating that the two groups diverged rather than improving in parallel. By day 90, participants receiving TruBlk™ had improved by 25.26 points (−51.8%) compared with 13.24 points (−27.4%) in the placebo group (difference in mean change 12.02 points). At Day 90, FSS was 11.58 points lower with TruBlk™ than placebo (95% CI 7.53 to 15.63), a large effect (Cohen’s d = −1.13, t(98) = 5.67, Sidak-corrected p < 0.0001) (Figure 4).

**Figure 4.**
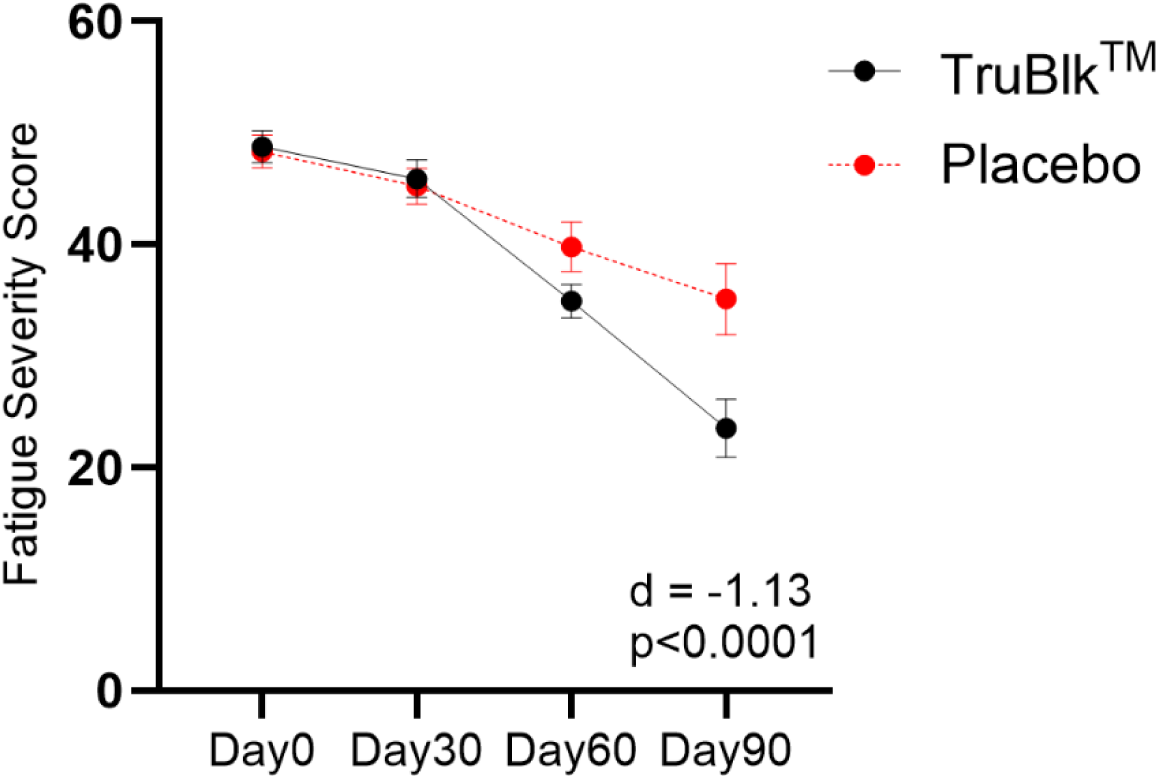
Fatigue Severity Scale over 90 days. FSS scores in participants receiving TruBlk™ (black, n = 50) or placebo (red, n = 50), assessed at baseline (Day 0) and at days 30, 60, and 90. Lower scores indicate reduced fatigue. Data are mean ± SEM.

#### 3.5.2 Participant and Investigator Global Assessment

At Day 90, Participant and Investigator Global Assessment scores were significantly more favourable in the TruBlk™ group than placebo (Participant: median Grade 2 [Much Improved] vs. Grade 4 [No Change], Mann-Whitney U p<0.0001; Investigator: median Grade 2 vs. Grade 4, p<0.0001). Using a responder threshold of "Much Improved" or "Very Much Improved" (Grades 1–2), 56% of participants in the TruBlk™ group met this threshold on self-rating compared with 14% in placebo (Figure 5A).

**Figure 5.**
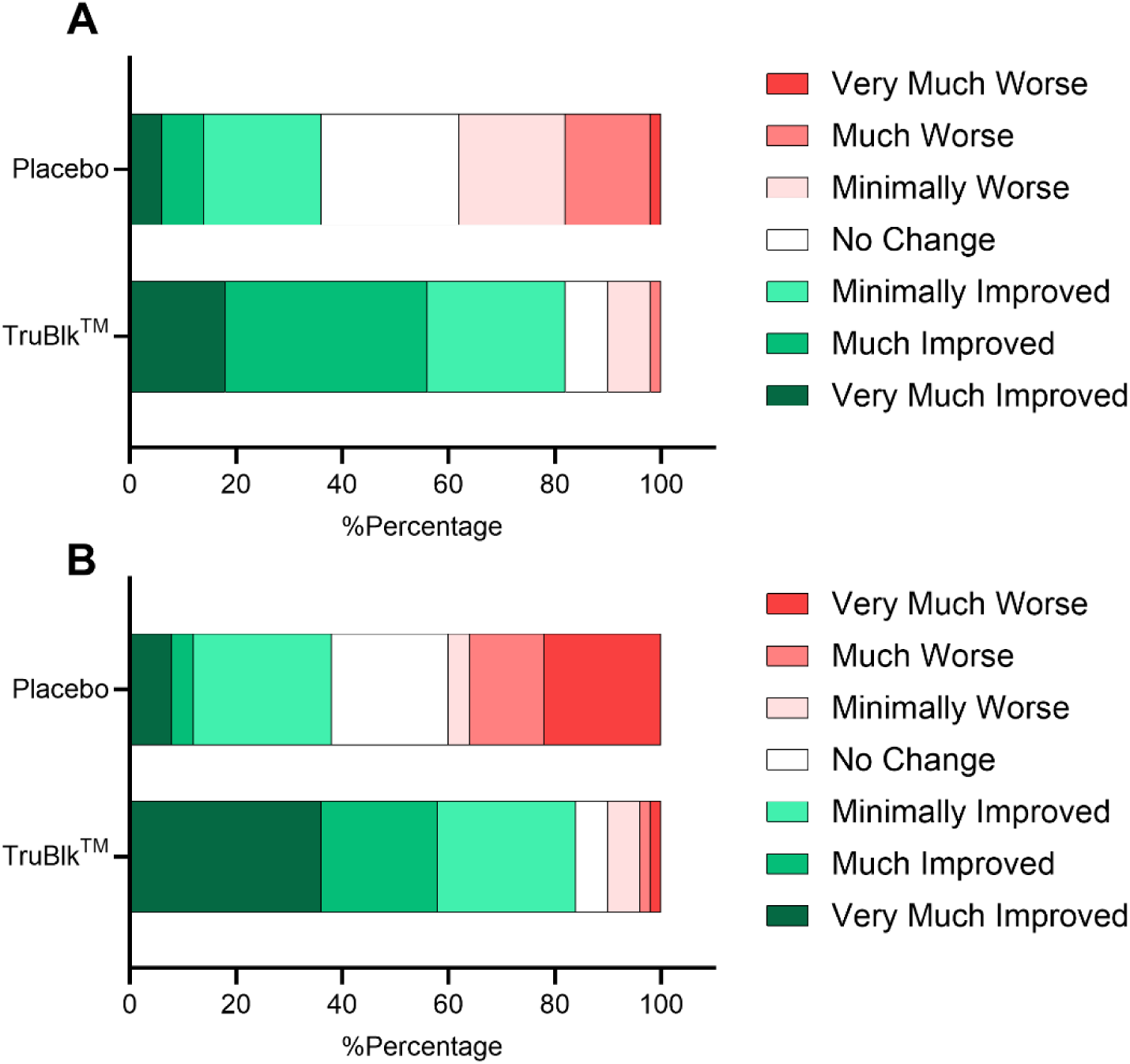
Participant and Investigator Global Assessment of overall improvement at Day 90. Distribution of response grades for (A) Participant Global Assessment (PGA) and (B) Investigator Global Assessment (IGA) in participants receiving TruBlk™ (n = 50) or placebo (n = 50), rated on a seven-point scale ranging from Very Much Improved to Very Much Worse. Bars show the percentage of participants in each grade.

Investigator ratings showed a closely similar pattern, with 58% of the TruBlk™ group rated "Much" or "Very Much Improved" compared with 12% in placebo. Using a broader threshold of any improvement (Grades 1–3), 82% of the TruBlk™ group and 36% of placebo showed improvement on participant rating, with a similar pattern on investigator rating (84% vs. 38%) (Figure 5B).

#### 3.5.3 Vascular Function

Nitric oxide was comparable between groups at baseline (median 24.55 [IQR 18.36–37.47] µmol/L in the TruBlk™ group vs. 23.48 [13.23–32.35] µmol/L in placebo; p = 0.157). Over the 90-day period, nitric oxide increased significantly within the TruBlk™ group (+7.43 ± 20.73 µmol/L, +25.7%; within-group p = 0.037), whereas the increase in placebo did not reach significance (+1.88 ± 22.12 µmol/L, +7.5%; p = 0.564). However, the between-group difference in change from baseline was not statistically significant (mean difference 5.55 µmol/L; Cliff’s δ = 0.08, p = 0.496).

### 3.6 Semen Parameters

Semen parameters were assessed in the subset of participants who consented to semen analysis (n = 57; TruBlk™ n = 29, placebo n = 28). Sperm concentration was comparable between groups at baseline (median 21.00 [18.00–25.00] vs. 20.00 [15.00–30.00] million/mL; p = 0.537) and increased from baseline with TruBlk™ (median change +4.00 [0.00 to 5.00] million/mL; within-group p < 0.001) but not with placebo (−2.00 [−3.00 to 2.00] million/mL; p = 0.255), a significant between-group difference (Cliff’s δ = 0.53, p < 0.001). Groups were also comparable at baseline for all motility parameters (p ≥ 0.078).

Total motility increased with TruBlk™ (median change +5.00 [3.75 to 11.00] percentage points; within-group p < 0.001) and was unchanged with placebo (−1.00 [−5.00 to 2.75]; p = 0.401), a significant between-group difference (Cliff’s δ = 0.63, p < 0.001). Progressive motility rose by 4.35 ± 5.48 percentage points with TruBlk™ (+10.9%; within-group p = 0.002) compared with −1.05 ± 7.57 (−2.8%; p = 0.543) with placebo, a between-group difference of 5.40 percentage points (95% CI 1.17 to 9.63; Cohen’s d = 0.82, p = 0.014). Rapid progressive motility also improved with TruBlk™ relative to placebo (median change +5.00 [0.00 to 5.00] vs. 0.00 [−3.50 to 0.00] percentage points; Cliff’s δ = 0.57, p = 0.001), whereas slow progressive and non-progressive motility did not differ between groups (p = 0.646 and p = 0.434)) (Table 4, Figure 6).

**Figure 6.**
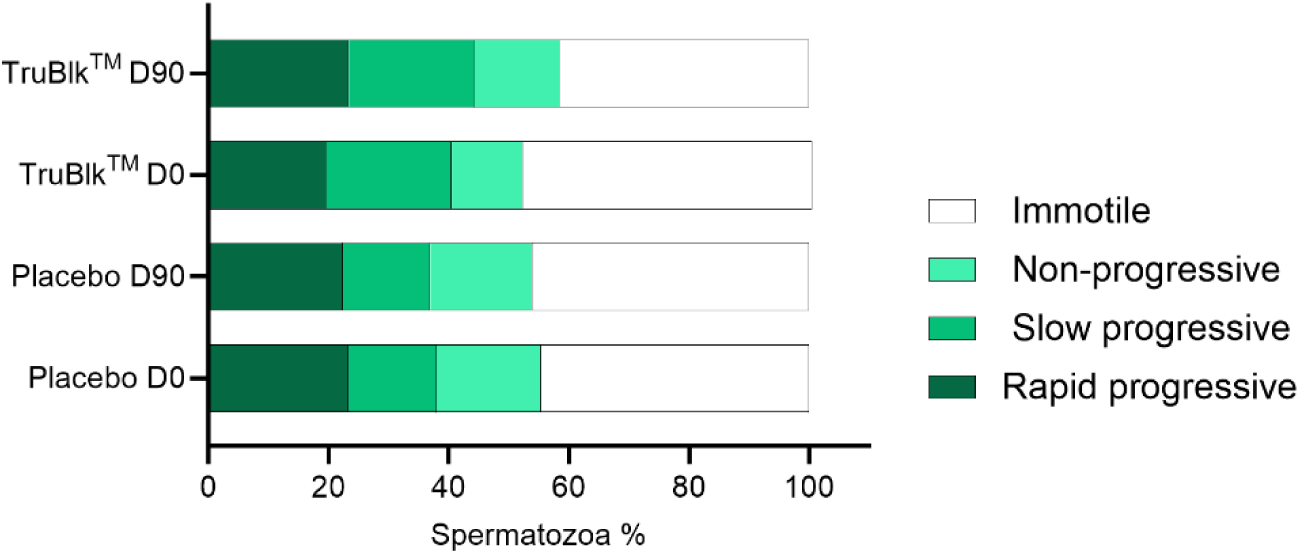
Distribution of sperm motility grades at baseline and Day 90. Mean percentage of spermatozoa classified as rapid progressive, slow progressive, non-progressive, or immotile in participants receiving TruBlk™ (n = 20) or placebo (n = 20) at baseline (D0) and Day 90 (D90).

**Table 4.** Semen Parameters at Baseline and Day 90.

| **Parameter** | **Baseline (TruBlk™ vs. Placebo)** | **TruBlk™ change** | **Placebo change** | **Between-group diff** | **Effect size** | **p** |
|---|---|---|---|---|---|---|
| Sperm concentration (million/mL)<sup>a</sup> | 21.00 [18.00–25.00]<br>vs.<br>20.00 [15.00–30.00], p=0.537 | +4.00 [0.00 to 5.00] | −2.00 [−3.00 to 2.00] | +3.00<br>(95% CI 2.00 to 6.00) | δ=0.53 | <0.001 |
| Total motility (%)<sup>b</sup> | 52.50 [30.00–70.00]<br>vs.<br>62.50 [43.75–70.50], p=0.615 | +5.00 [3.75 to 11.00] | −1.00 [−5.00 to 2.75] | +8.00<br>(95% CI 5.00 to 10.00) | δ=0.63 | <0.001 |
| Progressive motility (%)<sup>b</sup>† | 40.00±19.13<br>vs.<br>38.00±17.95, p=0.735 | +4.35±5.48<br>(+10.9%) | −1.05±7.57<br>(−2.8%) | +5.40<br>(95% CI 1.17 to 9.63) | d=0.82 | 0.014 |
| Rapid progressive motility (%)<sup>b</sup> | 19.50±10.25<br>vs.<br>23.25±13.11, p=0.320 | +5.00 [0.00 to 5.00] | 0.00 [−3.50 to 0.00] | +5.00<br>(95% CI 0.00 to 5.00) | δ=0.57 | 0.001 |
| Slow progressive motility (%)<sup>b</sup> | 15.00 [10.00–30.00]<br>vs.<br>15.00 [10.00–20.00], p=0.248 | 0.00 [−5.00 to 5.00] | 0.00 [−5.00 to 5.00] | 0.00<br>(95% CI −3.00 to 5.00) | δ=0.09 | 0.646 |
| Non-progressive motility (%)<sup>b</sup> | 10.00 [5.00–15.00]<br>vs.<br>15.00 [10.00–21.25], p=0.078 | 0.00 [0.00 to 5.00] | 0.00 [−3.50 to 5.00] | 0.00<br>(95% CI −3.00 to 5.00) | δ=0.14 | 0.434 |

*Values are mean ± SD with Cohen's d (unpaired t-test) where normally distributed, or median [IQR] with Cliff's δ (Mann-Whitney U test) otherwise.*

## 4. Discussion

This randomised, double-blind, placebo-controlled, multicenter trial evaluated standardised TruBlk™ Shilajit resin over 90 days in men aged 35–55 years with mild-to-moderate erectile dysfunction. TruBlk™ produced a significant improvement in IIEF total score, a primary endpoint, with a moderate effect size (Cohen’s d = 0.67), driven by significant gains in four of the five IIEF domains — erectile function, orgasmic function, overall satisfaction, and intercourse satisfaction — while sexual desire showed a smaller, non-significant improvement. This was accompanied by significant increases in total and free testosterone, with small-to-moderate effect sizes and no change in cortisol, and by significant Day 90 improvements in the sexual, energy and sleep domains of the HIS-Q. Among secondary endpoints, fatigue decreased markedly with TruBlk™ (FSS, large effect), and the improvements were reflected in how participants and investigators judged overall change: at Day 90, 56% of participants and 58% of investigators rated the TruBlk™ group as "Much" or "Very Much Improved," compared with only 12–14% for placebo. By contrast, nitric oxide showed no significant between-group difference. In the exploratory semen-analysis subset, sperm concentration and motility improved significantly with TruBlk™ relative to placebo.

The improvement in IIEF total score is consistent with, although not shown here to be mediated by, the well-established role of testosterone in erectile physiology. Androgens regulate the nitric oxide synthase (NOS)/cyclic guanosine monophosphate (cGMP) pathway within the corpus cavernosum, the principal mediator of cavernosal smooth muscle relaxation during erection, and preclinical work shows that androgen deprivation reduces cavernosal NOS expression and smooth muscle content while impairing erectile response, effects reversible with testosterone replacement [17, 18]. Testosterone additionally acts centrally in the regulation of sexual motivation and has been shown to preserve the structural integrity of cavernosal nerve fibres [19, 20]. Four of the five IIEF domains improved to a broadly similar, moderate degree, consistent with, but not demonstrative of, a hormonally mediated effect extending beyond erectile function alone; sexual desire was the exception, showing a smaller and non-significant change, which may reflect the more complex, less directly androgen-responsive nature of desire relative to the physiological steps involved in achieving and maintaining an erection.

The magnitude of these effects should be interpreted against realistic expectations. An oral supplement presumed to act through a slower, hormonally mediated pathway would not be expected to match the direct, rapid vasoactive effect of a phosphodiesterase-5 inhibitor such as sildenafil, and the moderate effect sizes observed here (d = 0.44–0.67) are in keeping with that expectation.

A further consideration is the substantial improvement observed in the placebo group across nearly every domain. Placebo response rates in erectile dysfunction trials are well documented to be unusually high, reported in phosphodiesterase-5 inhibitor trials at rates as high as 50%, and placebo responses in this condition may be accompanied by measurable physiological changes rather than reflecting subjective reporting alone [21]. Plausible contributors include the expectation and motivation associated with trial enrolment, increased attention from investigators and structured follow-up, and the act of monitoring and reflecting on sexual function through repeated questionnaire completion, all of which are well recognised in this literature independent of any active treatment. The improvement seen in the placebo arm of this trial is consistent with this broader pattern and should be read as expected background movement against which the TruBlk™ effect is measured, rather than as an anomaly.

The relationship between testosterone and fatigue is less settled in the peer-reviewed literature than commonly assumed. While decreased vitality is a commonly reported symptom in androgen deficiency, the Testosterone Trials’ dedicated Vitality Trial, a large, placebo-controlled RCT using a validated fatigue instrument in older hypogonadal men, found that testosterone treatment did not produce a clinically meaningful improvement in vitality relative to placebo [22, 23]. This tempers any straightforward causal reading of the FSS improvement observed here as testosterone-driven. A plausible alternative or complementary mechanism, inferred from preclinical work rather than demonstrated in this cohort, lies in shilajit’s proposed action on cellular energy metabolism. Fulvic acid and dibenzo-α-pyrones have been proposed to act as electron shuttles within the mitochondrial electron transport chain, and purified shilajit has been shown to increase ATP availability via improved mitochondrial function in preclinical models, a pathway independent of androgen status that this trial did not directly assess [24]. The HIS-Q Energy and Sleep domains were consistent with the FSS finding, both showing significant Day 90 differences favouring TruBlk™, neither of which is directly addressed by either the testosterone or mitochondrial-fatigue mechanisms discussed above. The Cognition and Mood domains, by contrast, did not reach significance at Day 90 despite significant treatment × time interactions, suggesting the improvement may be more specifically tied to energy and sleep, which share closer physiological overlap with fatigue, than to the broader symptom complex of androgen deficiency as a whole.

Shilarathenes™ also comprises a spectrum of urolithin-associated metabolites and phenolic acids, and individual compounds within both classes have reported biological activity relevant to the outcomes measured here, although their contribution to the effects of the TruBlk™ preparation was not assessed. Urolithins have been shown to increase NO production and endothelial nitric oxide synthase expression in human endothelial cells [25], with a placebo-controlled human trial demonstrating improved vascular endothelial function following urolithin-A supplementation, albeit dependent on gut microbiota [26]. Phenolic acids, separately, have been shown to inhibit phosphodiesterase-5 directly in penile tissue in preclinical models [27], and a meta-analysis of clinical trials found that Pycnogenol®, a pine bark extract rich in procyanidins and phenolic acids, improved erectile function when given with L-arginine [28]. While this trial did not isolate the contribution of individual shilarathenes™ constituents, it is plausible, though untested here, that more than one bioactive fraction contributes, rather than a single mechanism operating alone.

Notably, nitric oxide, a marker often associated with vascular and endothelial function, showed no significant between-group difference despite the improvement in IIEF total score. This dissociation suggests that the improvement in erectile function observed here may not be primarily driven by improved systemic vascular nitric oxide bioavailability, and is compatible with an androgen-mediated contribution, although this trial did not test whether the testosterone changes mediated the IIEF improvement. This interpretation warrants a caveat: nitric oxide was measured here as a circulating serum marker, reflecting systemic endothelial NO production rather than local penile haemodynamics or cavernosal tissue NOS activity specifically [29]. Testosterone is known to regulate NOS expression directly within cavernosal tissue through local mechanisms that a systemic serum marker would not necessarily capture, so this trial cannot fully exclude a localised vascular contribution to the erectile function improvement observed.

In the semen subset, sperm concentration increased significantly with TruBlk™ relative to placebo, and total, progressive, and rapid progressive motility improved in parallel. These findings are consistent with a prior trial of processed Shilajit in men with diagnosed oligospermia, which reported increases in total sperm count and motility alongside higher serum testosterone and lower seminal malondialdehyde over the same 90-day duration [30], and with rodent studies in which Shilajit increased sperm count and testicular daily sperm production [31, 32]. Two mechanisms may plausibly contribute, though neither was tested here. Spermatogenesis is androgen-dependent [33], so the rise in testosterone over an intervention longer than one spermatogenic cycle [34] could support sperm output. Separately, sperm motility relies on mitochondrial ATP production and is readily impaired by oxidative stress [35, 36], which the antioxidant and mitochondrial actions proposed for fulvic acid and dibenzo-α-pyrones [24] could counter. As the changes were modest and the motility analysis was restricted to one site and not pre-specified, these findings should be regarded as exploratory and do not establish any effect on fertility or pregnancy outcomes.

In conclusion, ninety days of standardised TruBlk™ Shilajit resin produced improvement in erectile function and several domains of the broader hypogonadism-related symptom complex in men with mild-to-moderate erectile dysfunction, alongside favourable shifts in testosterone and fatigue, and exploratory evidence of improved sperm concentration and motility. These results provide a randomised, placebo-controlled, multicenter evidence that standardised Shilajit resin improves sexual function in a clinically diagnosed population, rather than the healthy-volunteer cohorts studied to date, and establish a rationale for the mechanistic and larger-scale studies outlined above.

## Data Availability

All data produced in the present study are available upon reasonable request to the authors

